# Dynamic Simulation Modeling to Analyze the Impact of Whole Genome Sequencing National Implementation Scenarios in Lung cancer on Time-to-Treatment, Costs and Patient Demand

**DOI:** 10.1101/2023.11.10.23298371

**Authors:** Michiel van de Ven, Hendrik Koffijberg, Valesca P.J. Retèl, Wim H. van Harten, Maarten J. IJzerman

## Abstract

**Background:** Although Whole Genome Sequencing (WGS) is increasingly proposed to unravel molecular origins of advanced cancers, it is less clear if and how WGS should be routinely offered in the health service. The objective of this study is to investigate how the cost per patient and time-to-treatment is affected if WGS were implemented in the national health system and how these outcomes differ among subgroups of patients with lung cancer. This first-ever study used health systems simulation modeling to analyze implementation scenarios ensuring sustainable access to cancer treatment.

**Methods:** A base case and three scenarios (varying stage of disease and hospitals offering WGS) the optimal placement of WGS in the diagnostic pathway was simulated using a dynamic simulation model. The model simulated lung cancer patients undergoing molecular diagnostic procedures in one or multiple hospitals. The model also included patient and healthcare provider heterogeneity as well as referral patterns of lung cancer (LC) patients using patient-level data obtained from the Netherlands Cancer Registry. Model outcomes were the time-to-treatment, total diagnostic cost, and the demand for WGS sequencing capacity including the expertise of a molecular tumor board.

**Results:** The time-to-treatment ranged between 20-46 days for all four scenarios considered. The cost of molecular diagnostic testing per patient ranged from €621 in the base case to €1930 in the scenario where all LC patients (stage I-IV) receive upfront WGS. Compared to the base case, upfront testing using WGS in all LC patients led to a 33% reduction in the time-to-treatment, a 210% increase in the cost per patient and a six-fold increase in total diagnostic costs.

**Conclusions:** This first-ever study investigating implementation scenario’s demonstrated that upfront WGS for all lung cancer patients can reduce the time to treatment yet at a higher cost. However, upfront WGS also reduces diagnostic pathway complexity, which may improve care planning and treatment efficiency. The model is versatile in its approach to study the impact of price discounts or the amount of actionable targets tested for and further analysis showed discounts on consumables up to 50% imply WGS would the preferred strategy.

## 1. Introduction

Whole Genome Sequencing (WGS) is a comprehensive genomic test that analyzes the entire genome and therefore delivers a higher diagnostic yield compared to the commonly used targeted single gene tests or targeted gene panels. Several countries have taken initiatives for the implementation of WGS in clinical oncology [1], with an emphasis on rare cancers and cancers with unmet needs.

There are multiple areas in which WGS can provide additional value compared to the current standard of care (SOC) for treatment selection, in particular in assisting with diagnosing complex or rare tumor types such as carcinoma of unknown primary (CUP) and blood cancers [2, 3]. Furthermore, WGS is currently used as a last-resort diagnostic for patients with refractory cancers to identify actionable mutations, which would otherwise be complex and inefficient to identify using SOC testing. Moreover, WGS can be considered a substitute in cancers where multiple SOC tests are required to test for all relevant biomarkers. WGS can increase the efficiency of the diagnostic pathway by streamlining workflows [4]. Ultimately, this may have benefits for the healthcare system and for patients. Implementation studies such as “WGS Implementation in the standard Diagnostics for Every cancer patient” (WIDE) study in the Netherlands, aim to investigate the feasibility to provide WGS-based diagnostics as part of routine diagnostics for metastatic cancer patients [5].

While the clinical evidence for using WGS is increasing, it is less clear how the benefits are to be demonstrated. First, it is not defined which patient subgroups would benefit most from upfront WGS and how that will change over time when (more) evidence of clinical utility becomes available. Second, the extent that current SOC testing will be replaced, i.e., the degree of substitution, by WGS is unclear. Tumor types such as lung cancer require multiple different biomarker tests conducted for a clinical diagnosis, such as immunohistochemistry, Next Generation Sequencing (NGS) panels, and fluorescence in situ hybridization [6]. While WGS can be a substitute for all DNA-based biomarker tests, the most efficient use of WGS as either an upfront test for all or for metastatic cancers only is unknown. Third, realizing the benefits of WGS implies that there should be equal access for patients and, hence, enough hospitals need to offer WGS and associated treatments. Performing a prospective clinical study would be challenging and only partly provide the answers to these questions, so a simulation model that can analyze different scenarios would be appropriate ahead of implementation.

In addition to the modeling that is used to inform reimbursement decisions, simulation models can potentially inform health policy by incorporating implementation characteristics relevant for actual use in the health service. This particularly is relevant as WGS is a disruptive technology and thus the full benefits of WGS can only be realized if care pathways are adapted to accommodate WGS. For example, data curation and clinical interpretation of WGS data should be performed in molecular tumor boards (MTB) [7,8], which are typically not needed for SOC testing.

Simulation models can be implemented using Dynamic Simulation Modeling (DSM), which is a set of modeling approaches. Amongst others, DSM can be used to model patient-level variation, care provider heterogeneity, dynamic diagnostic and treatment processes [9,10] and can reflect the multi-levelled nature of macro-level or nationwide implementation of WGS.

The primary objective of this study is to investigate how the cost per patient and time-to-treatment is affected by the nationwide implementation of WGS as a cancer diagnostic and how these outcomes differ among patient subgroups. Both current costs of WGS and discounted costs of consumables for WGS will be used. This discount is potentially attainable when conducting WGS at scale. The secondary objective is to estimate the aggregate demand for sequencing and analytic capacity with the respect to WGS, based on the assumed delivery of WGS services.

In the analysis, a base case reflecting the current situation and three scenarios will be analyzed: one with a 2-year time horizon and two with a 5-year time horizon. These scenarios differ on three dimensions: the patient indication, the hospital types that use WGS, and the position of WGS in the diagnostic strategy. The scenarios will be simulated with a previously developed dynamic simulation model [11] that reflects the organization of care for WGS in the Netherlands. Non-small cell lung cancer (NSCLC), the largest subtype of lung cancer, will be used a case study, as the role of molecular diagnostics is substantial in the prediction of treatment response in this cancer type [12].

## 2. Methods

### 2.1 Health Systems Simulation model

The previously created dynamic simulation model [11] on the diagnostic pathway for NSCLC allows studies into the affordability and accessibility of the use of WGS. The model reflects the diagnostic pathway for lung cancer in the Netherlands and included patient and institute heterogeneity, diagnostic workflows, referral patterns, and a spatial representation of the hospital landscape in the Netherlands. For a detailed technical description of the simulation model and input parameters, we refer to the supplementary document of van de Ven et al. [11]. The simulation model can be inspected and run on AnyLogic Cloud [13].

### 2.2 Outline of the simulation

Figure 1 provides a schematic representation of the model structure. The model was implemented as a hybrid dynamic simulation model, combining Discrete-Event Simulation (DES) and Agent-Based Modeling (ABM), and was developed in AnyLogic 8.3.3 (The AnyLogic Company). The model contained the following agents: academic (n = 8), teaching (n = 21), and general (n = 43) hospitals, a sequencing facility (n = 1), regional MTBs (n = 8). Additionally, patients with NSCLC were being generated over time according the to the incidence of NSCLC patients in the Netherlands [14].

**Figure 1.**
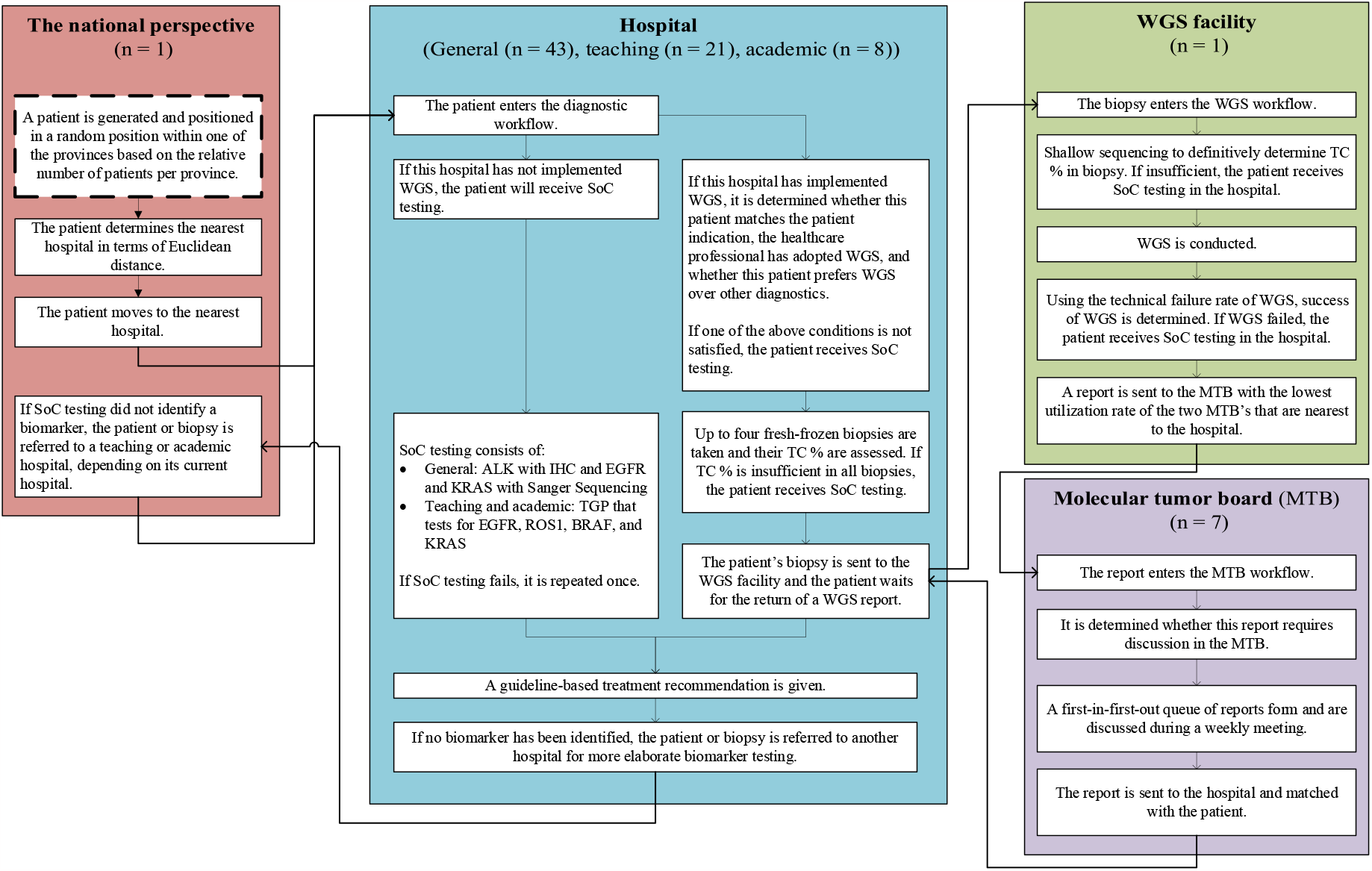
Schematic representation of the model structure, adapted from [11]. SoC, Standard of Care; WGS, Whole Genome Sequencing; IHC, immunohistochemistry; TGP, targeted gene panel; TC, tumor cell; MTB, molecular tumor board. The flow of the figure starts at the box with the dashed outline in the national perspective

Hypothetical patients with NSCLC who require biomarker testing for initial treatment selection are generated and enter the diagnostic workflow of the nearest hospital to receive biomarker testing, either a form of SOC testing or WGS. Depending on the scenario, WGS is preceded by one or multiple tests. If the patient receives WGS, the biopsy material is sent to the WGS facility, and if sequencing was successful, a report is sent to the MTB nearest to the hospital for a clinical interpretation. The biopsies of patients are referred to another hospital with more extensive testing capabilities if no actionable target has been identified so far and one or more biomarkers has not yet been tested. The selection for the referral hospital is dependent on distance and on the type of the referring hospital, so that a general hospital refers to nearest teaching hospital, and a teaching hospital refers to the nearest academic hospital. The endpoint in the model is when either SOC has been concluded or when the clinical interpretation of WGS results have been reported back to the hospital.

### 2.3 Base case and simulated implementation scenarios

In the base case, all patients receive SOC. Three implementation scenarios were defined in consultation with a medical oncologist and the managing director of the Hartwig Medical Foundation, the one central facility in the Netherlands that conducts WGS for cancer patients that are enrolled in clinical trials. The scenarios describe potential variants of how WGS might be used as a diagnostic test for lung cancer in the Dutch healthcare system in the future. This includes a scenario with a two-year time horizon (short term), and two scenarios with a five-year time horizon (long term). To provide a bandwidth of possible outcomes, these two longer-term scenarios describe a neutral and a progressive perspective regarding the implementation rate of WGS. Table 1 provides a summary description of the content of the base case and scenarios.

**Table 1.** Characteristics of the implementation scenarios as well as the current situation (base case). IHC: immunohistochemistry; PD-L1: programmed death-ligand 1.

| Scenario | Base case | New scenario's for use of WGS in |  |  |
| --- | --- | --- | --- | --- |
|  |  | <2 years | 5 years<br>(conservative) | 5 years<br>(progressive) |
| **Patient indication** | Only SoC is used for all patients | Only stage IV NSCLC patients receive WGS (n = 5313/year [15]) | Only stage III and IV NSCLC patients (n=7550/year [15]) | All lung cancer patients (n = 9974/year [14]) |
| **Hospital offering WGS** | WGS is used in none of the hospitals | Only academic hospitals and not teaching or community hospitals |  | All hospitals (Academic, teaching, community) |
| **Where WGS is used in the molecular diagnostic pathway** | Only SOC diagnostics are used | WGS after an IHC test for PD-L1 | WGS after a NGS panel and IHC test for PD-L1 | WGS upfront in all patients |

The implementation scenarios vary across three dimensions: the patients for whom WGS is used, the type of hospitals offering WGS, and the position of WGS in the biomarker test strategy. In scenarios that state that only academic hospital services use WGS, only patients that are receiving diagnostics in one of the academic hospitals can receive WGS, and patients who are receiving diagnostics in either a teaching or general hospital will only be able to receive SOC, after which they may be referred to an academic hospital if necessary. The cost of consumables make up the majority of the total cost of WGS at € 2187 of a total of € 2925 per patient [16]. Suppliers may be able to give discounts when conducting WGS at scale. To illustrate the effects of these potential discounts, we also simulated the base case and the scenarios using a reduced cost of WGS, in which the cost of consumables was decreased by 50%. This reduction in the cost of consumables reduced the cost of WGS to €1831 per patient (see supplement, figure S1).

#### 2.3.1 Outcomes

The outcomes of interest are the time-to-treatment (TTT), costs per patient for biomarker testing (€), total annual diagnostic cost (€), and the aggregate demand for WGS (N). The time-to-treatment is defined as the time in days from initiating the first biomarker test until the start of treatment. The aggregate demand for WGS is split into the number of biopsies that enter the WGS workflow and the number of WGS reports that require a clinical interpretation in MTBs. The average annual diagnostic cost is the sum of the costs for all patients across all hospitals, divided by the number of years in simulation time. Outcomes are stratified by patient subgroups: patients for whom WGS was not initiated, patients for whom WGS was successfully completed, and patients for whom WGS was initiated but not successfully completed. Not being able to complete WGS successfully can be due to not meeting the tumor percentage requirements or due to technical failures. Not initiating WGS can be due to not matching the patient indication, receiving only SOC as WGS was not used in that specific hospital, patient preferences, or due to a healthcare professional that has not adopted WGS. If WGS was not initiated for patients, they received SOC diagnostics.

### 2.4 Analysis

For the base case and each scenario, the simulation model was run 1000 times to quantify the stochastic uncertainty in the outcomes [17]. Each simulation ran for 2500 days, but the generation of new patients halted on day 2000 to ensure that enough patients were simulated while also giving all patients opportunity to flow through the model. The simulated patient-level outcomes accumulated by patients generated between 500 and 2000 days were used in the analysis. The data analysis and visualization was conducted using R software version 4.0.3 [18].

## 3. Results

### 3.1 Time-to-treatment

Figure 2 shows the distribution of the time-to-treatment for patients in each scenario. Comparing scenario ‘long term (progressive)’ with the other scenarios, figure 2 shows that the mean time to treatment is shorter for patients in scenario ‘long term (progressive)’ compared with the other scenarios (base case: 30 days, short term: 43 days, long term (neutral): 46 days, long term (progressive): 20 days). This is because biopsies of patients are not referred to another hospital if WGS was initiated, as there is no superior test available in other hospitals. In scenario ‘long term (progressive)’ all patients are eligible for WGS, but WGS is not initiated for all patients, as we assumed that 90% of healthcare professionals has adopted WGS and 90% of patients prefer WGS over other diagnostics. Moreover, biopsies of patients for whom WGS was not initiated may still be referred if SOC testing identified no actionable biomarker.

**Figure 2.**
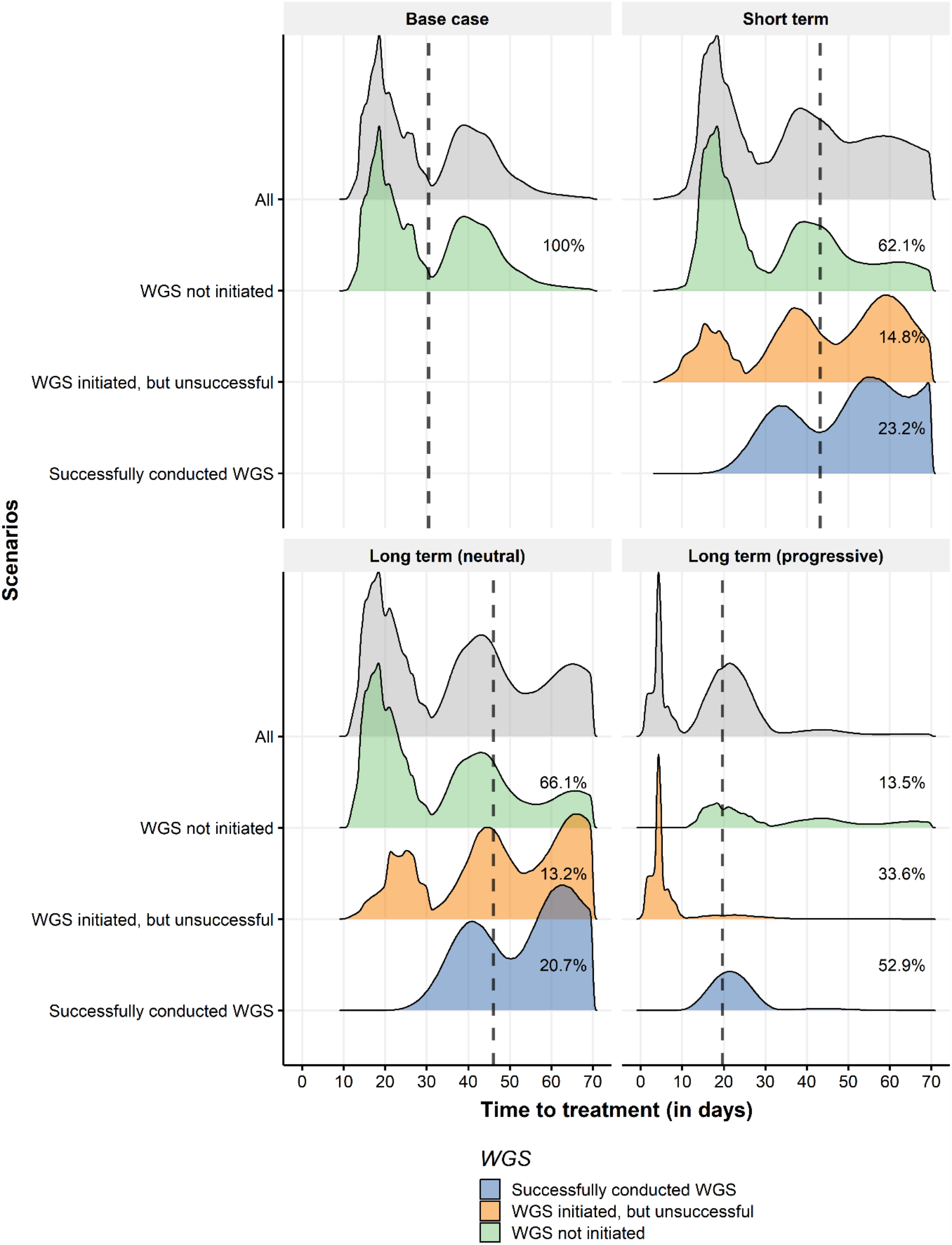
Distribution of the time-to-treatment for the base case and three scenarios. Subgroups reflect patients for whom WGS was not initiated (green), WGS was initiated but not completed successfully (orange), and WGS was successfully conducted (blue). Grey density curves are all these subgroups combined. The dashed line illustrates the mean time-to-treatment in each scenario across all subgroups. The percentages shown for each subgroup represent the size of each subgroup. The area under the density curves for each subgroup is reflective of the percentage of patients in each subgroup for each scenario. Patients who did not conclude their diagnostic pathway are not included in this figure.

To explain the peaks in the figure, looking at scenario ‘short term’, the first peak at 20 days for the subgroup of patients for whom WGS was not initiated, represents a group of patients for whom their biopsy was not referred to another hospital as an actionable biomarker was identified. The second peak at 40 days represents a group of patients for whom their biopsy was referred once either to a teaching or academic hospital. The remainder of patients with a time-to-treatment beyond 50 days are patients for whom their biopsy was sent to two other hospitals, as SOC testing in the general and teaching hospital were unable to identify an actionable target, and thus, these biopsies were referred finally to an academic hospital. Additionally, if the initial attempt of SOC testing fails, SOC testing is repeated once, which increases the time-to-treatment.

### 3.2 Cost per patient

The mean cost per patient was € 621 in the base case, € 1444 in the ‘short term’, € 1495 euros in the ‘long term (conservative)’, and € 1930 in ‘long term (progressive)’ scenario. The mean cost per patient for patients in whom WGS was successfully conducted in scenario ‘long term (progressive)’ is substantially lower compared to the same subgroup in other scenarios (€ 3595 in scenario ‘short term’, € 3877 in scenario ‘long term (conservative)’, € 2951 in scenario ‘long term (progressive)’. In scenario ‘long term (progressive)’, this patient subgroup only receives WGS and no prior SOC diagnostics.

The multiple peaks that are displayed in Figure 3 are caused by the different diagnostic trajectories that patients traversed in the simulation model. As the definition of SOC testing varies between hospital types, so do the costs that SOC testing incurs. This means that it matters to which hospital type a patient first presents themselves. Moreover, as biopsies of patients may be referred, they incur costs in multiple hospitals. Additionally, patients may receive SOC or WGS, or both, leading to differences in costs between patients. Furthermore, SOC may have a technical failure, in case which SOC is repeated once, and thus, costs for SOC are counted twice. Likewise, not all biopsies have a high enough tumor cell percentage to be used for WGS, which means that only the costs for shallow sequencing are included, which is 25% of the total costs for WGS.

**Figure 3.**
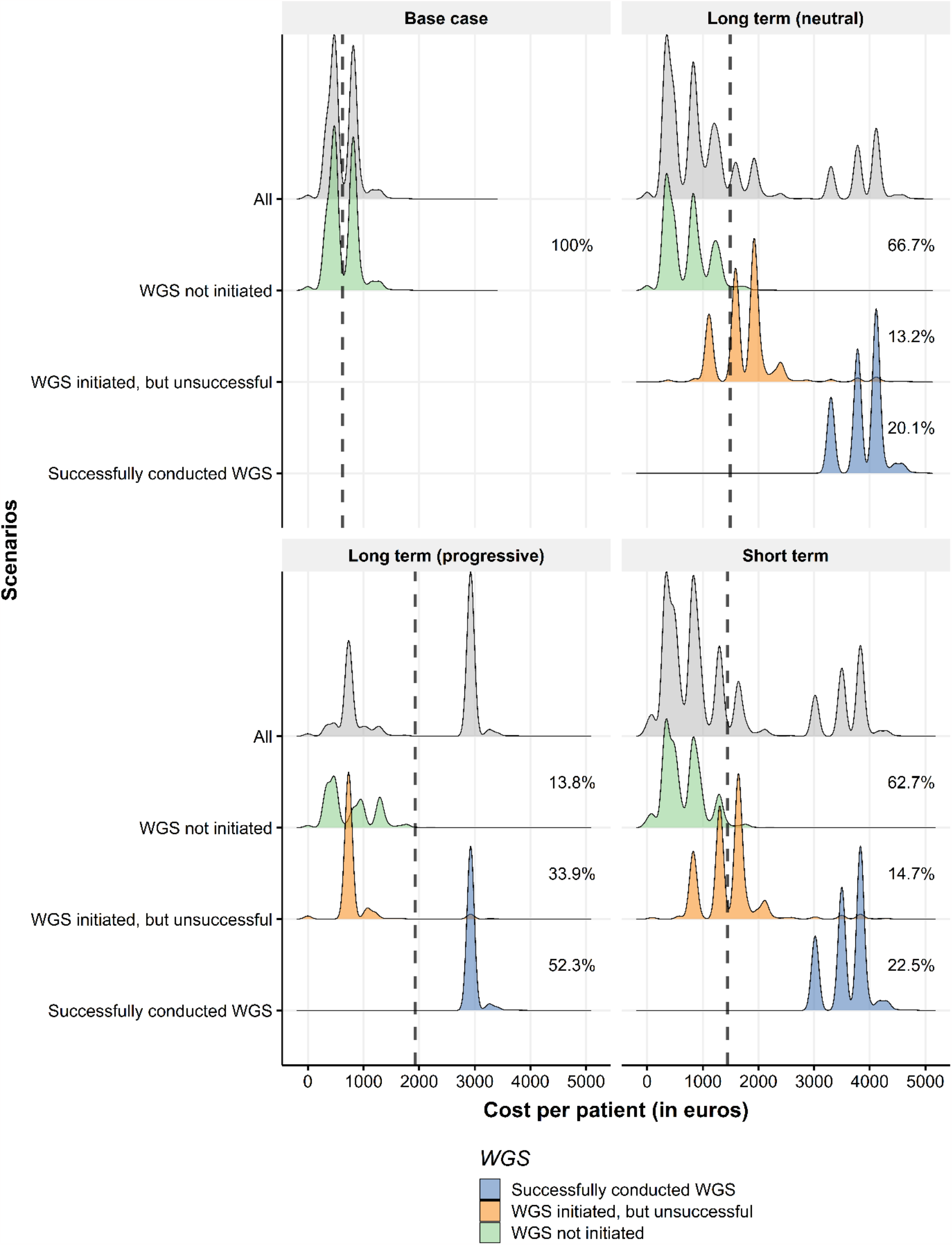
Distribution of the cost per patient for the base case and three scenarios. Subgroups reflect patient groups for whom WGS was not initiated (green), WGS was initiated but not completed successfully (orange), and WGS was successfully conducted (blue). Grey density curves are all these subgroups combined. The dashed line illustrates the mean cost per patient in each scenario across all subgroups. The percentages shown for each subgroup represent the size of each subgroup. The area under the density curves for each subgroup is reflective of the percentage of patients in each subgroup for each scenario.

The total annual diagnostic cost, averaged over all simulation runs, is € 4.2 million (SD: € 54,000) in the basecase, € 9.7 million (SD: € 96,000) for scenario ‘short term’, € 14.3 million (SD: € 119,000) for scenario ‘long term (neutral)’, and € 24.5 million (SD: € 148,000) for scenario ‘long term (progressive)’.

### 3.3 Discounted cost of consumables for WGS

If the cost of consumables is discounted by 50%, the cost of WGS would fall to 1831.65 euros. Figure S1 in supplementary file 1 shows that the mean cost per patient in scenario ‘long term (progressive)’ at 1258 euros is approximately equal to the mean cost per patient of scenario ‘long term (neutral)’ at 1236 euros, in which the use of and access to WGS is more limited. Compared to Figure 3 which uses the discounted cost of WGS, Figure S1 shows that the cost distribution for all subgroups for whom WGS was initiated is shifted to the left because of the lower cost of WGS. While scenario ‘long term (progressive)’ still has the highest mean cost per patient at 1258 euro per patient, this is below the mean cost per patient in scenarios ‘short term’, ‘long term (neutral)’, and ‘long term (progressive)’ shown in Figure 3 which uses the unreduced cost of WGS.

Using the reduced cost of WGS, the total annual diagnostic cost is 4.2 million euros (SD: 54,000 euros) in the base case, 7.8 million euros (SD: 73,000 euros) for scenario ‘short term’, 11.9 million euros (SD: 88,000 euros) for scenario ‘long term (neutral)’, and 15.9 million euros (SD: 100,000 euros) for scenario ‘long term (progressive)’.

### 3.4 Aggregate demand

Table 2 lists for each scenario the annual number of biopsies that were sent to the WGS facility for sequencing, the annual number of biopsies that passed quality control, and the annual number of successfully sequenced biopsies. Table 2 also shows the total number of reports that were sent to and discussed in MTBs, as well as the average number of reports received per MTB, as some MTBs receive more reports than other MTBs.

**Table 2.** The aggregate demand (number of biopsies processed) for WGS for each scenario. Outcomes are averaged across simulation runs. Means and Standard Deviation (SD) presented.

| Scenario | Biopsies sent for WGS per year | Biopsies passed quality control/year | Biopsies successfully sequenced per year | Reports received by all MTBs per year | Reports received per MTB per year |
| --- | --- | --- | --- | --- | --- |
| Base case | 0 | 0 | 0 | 0 | 0 |
| <2 years | 1773 (± 37) | 1688 (± 36) | 1647 (± 35) | 1647 (± 35) | 235 (± 61) |
| 5 years (conservative) | 2255 (± 47) | 2147 (± 45) | 2094 (± 44) | 2092 (± 45) | 299 (± 74) |
| 5 years (progressive) | 7549 (± 43) | 7194 (± 43) | 7040 (± 43) | 7044 (± 43) | 1006 (± 351) |

## 4. Discussion

In this study, we investigated how the cost per patient and time-to-treatment is affected by the nationwide implementation of WGS as a cancer diagnostic and how these outcomes differ among patient subgroups. Additionally, we estimated the aggregate demand for sequencing and analytic capacity with the respect to WGS. When WGS is used upfront for all stage I-IV lung cancer patients, the mean cost per patient (1930 euros) and total annual diagnostic costs (24.5 million euros) were highest while the mean time-to-treatment (20 days) was lowest, compared to the other scenarios. Also, the aggregate demand for WGS was highest when WGS was used upfront for all patients, because of the increased access to WGS and large patient indication. It should be noted that any decision on the use of WGS in a particular setting depends on the clinical utility or need.

Our results show that using an NGS panel prior to WGS makes little difference for the cost per patient and time-to-treatment. Scenario ‘short term’ only uses a PD-L1 test prior to WGS, scenario ‘long term (neutral)’ uses both a PD-L1 test and an NGS panel prior to WGS. Our simulation results show that including a PD-L1 test and NGS panel prior to WGS increases the time-to-treatment by three days at similar costs per patient. Therefore, based on our results, using both an NGS panel and WGS in sequence does not seem to efficient use of resources when considering the time-to-treatment and testing cost per patient.

The results indicate there is a tradeoff between costs and time-to-treatment across implementation scenarios. For example, the cost per patient and total annual diagnostic costs are lowest in the base case where WGS is not used at all, while the mean time-to-treatment is 10 days longer compared to a scenario where all patients are receiving upfront WGS, which has the shortest mean time-to-treatment and highest cost per patient and total annual diagnostic costs. On the one hand, the mean cost per patient for the subgroups for whom WGS was successfully conducted is substantially higher than for other patients. On the other hand, conducting WGS upfront in all patients such as in scenario ‘long term (progressive)’ means that biopsies of patients are no longer referred to another hospital as no other hospital has a more extensive diagnostic test available.

From an organizational perspective, upfront testing for all patients may also have other benefits, as diagnostic workflows can be simplified through the substitution of current SOC tests. Recently, it has been shown that conducting WGS once is sufficient for almost all patients to identify SOC biomarkers [19]. Besides consequences for the cost-effectiveness of WGS, this also means that the amount of biomarker tests needed if treatment progression or resistance is detected, is reduced for patients for whom WGS was already conducted. In effect, this increases the degree of substitution that WGS brings and further helps to simplify the diagnostic pathway. This also means that aggregated across multiple treatment lines, the testing costs with WGS upfront will become more favorable, relative to SOC testing costs, than reported here.

Conducting WGS at a higher volume may make it possible to achieve a lower cost per patient, by increasing the utilization rate of sequencing devices [20] and by receiving a volume discount on certain cost drivers of WGS. The likelihood of obtaining and the magnitude of a volume discount is partly dependent on the demand for WGS. It is therefore more likely that this discount can be obtained when the patient indication is largest, such as in the scenario in which WGS used upfront for all lung cancer patients and is less likely when the demand is more limited such as in the other scenarios.

In an additional analysis, we applied a 50% discount on the cost on consumables. This led to a mean cost per patient for scenario ‘long term (progressive)’ of 1258 euro (Figure S1), down from a mean cost per patient of 1930 euro (Figure 3). This cost of 1258 euro per patient is lower than the mean cost per patient in all scenarios when the original cost of WGS was used. Although a 50% discount is substantial, and may not (immediately) be possible, having a higher demand for WGS leads to an improved bargaining position with suppliers to receive a high volume discount. Thus, conducting WGS upfront in all patients may initially lead to a higher cost per patient and total annual diagnostic costs. However, in the end it may prove to be the approach with the shortest time-to-treatment and most inexpensive of all three implementation scenarios if a substantial volume discount is obtained.

On a more cautionary note, the time-to-treatment of 20 days that may be attained by conducting WGS upfront for all patients is only possible if enough sequencing and analytic capacity is available to meet WGS demand (Table 2). Otherwise, insufficient capacity may cause long delays in the diagnostic pathway to the point that patients may not wait for WGS results but rather start with a suboptimal treatment. Implementation and infrastructure building strategies can help to prevent such bottlenecks by deliberately focusing on genomic workforce education, which has been recognized widely as important for an optimal clinical translation of genomic data [21,22]. Additionally, putting in place infrastructure and tools to improve the efficiency of MTB’s, such as the use of clinical decision support systems is underway [23].

It is unlikely that WGS will fully and completely substitute current SOC diagnostics, considering that currently WGS cannot be successfully conducted for 28% of biopsies [24]. This is primarily due to not meeting the tumor cell percentage requirements. Thus, it is likely that SOC biomarker tests, and the corresponding infrastructure and logistics, will need to remain available. These can serve as alternatives in cases where WGS cannot be completed successfully.

To our knowledge, this study is the first systems modeling approach to implementing genomics in healthcare. The main strength of this study is that it quantifies the consequences of multiple implementation scenarios, addressing key uncertainties in the potential future use of WGS, while adopting a whole-systems perspective [25]. Hence, this study is able to go beyond mentioning the cost per patient in a single institute by estimating the time-to-treatment along the entire diagnostic pathway and the total diagnostic costs associated with each scenario. Combined with the estimates for aggregate demand, these outcomes can inform implementation and infrastructure building strategies to prepare the healthcare delivery system for increased use of WGS in oncology. And thus, our results have additional relevance for policy, which is not captured in other studies. This system modelling approach ideally complements more standard economic evaluation approaches estimating cost-effectiveness [26]. However, these models are increasingly complex because of the range of mutations and the follow-on treatment that significantly drives up the cost.

While a systems approach has several strengths, this study also has some limitations. First, the simulation is limited to the diagnostic pathway. Therefore, the consequences of the potentially improved treatment selection and the costs of treatments have not been included and therefore, the results are likely underestimations of the true societal benefit. It is likely that WGS has the potential to improve treatment selection by more accurately predicting which treatment will work best and by helping to prevent potentially ineffective treatment. Hence, it is plausible that increased use of WGS will indirectly lead to patient benefit.

The second limitation of this study is that the results cannot be directly generalized to other tumor streams. We chose to focus only on NSCLC as including other cancer types would lead to increased model complexity due to differences in patient populations and diagnostic pathways. However, NSCLC is a relevant case study as it can be used to illustrate the substitution effects of WGS and has a high incidence. Consequently, how WGS is used for this cancer type can substantially affect the total demand for WGS across all cancer types.

Third, differences in diagnostic yield across biomarker tests were not incorporated, and thus, the study implicitly assumes equal diagnostic yield across all tests. Incorporating differences in diagnostic yield could potentially lead to increased subgroup differences.

There are multiple interesting avenues for future research. A further study could investigate which scenario would be most desirable, creating a target for implementation and infrastructure building strategies. Additionally, it needs to be determined which actions are required to realize that scenario.

## Data Availability

All data produced are available online at https://cloud.anylogic.com/model/6f5c67f2-1423-422a-be35-63f0f664cc77?mode=SETTINGS

## Acknowledgements

This study was financed by a grant from ZonMW (grant number 80-84600-98-1002). ZonMW otherwise was not involved in this study. This study is created in collaboration with the Technology Assessment of Next Generation sequencing for personalized Oncology (TANGO) consortium. The experts employed by the Hartwig Medical Foundation that were consulted for the definition of the implementation scenarios had no involvement in the study design, objective, analysis, interpretation, and drafting the manuscript for this study.

## Author contributions

Conceptualization: MV, HK, MIJ; Writing manuscript: MV, MIJ; Formal modeling and analysis: HK, MV; Visualization: MV, HK, MIJ; Review & editing: MV, VR, HK, WH, MIJ; Funding: VR, WH, MIJ.

## Ethics Declaration

The need for ethics approval is waived, as the study does not concern medical scientific research and does not include human subjects.^24^

## Supplementary information

**Figure S1.**
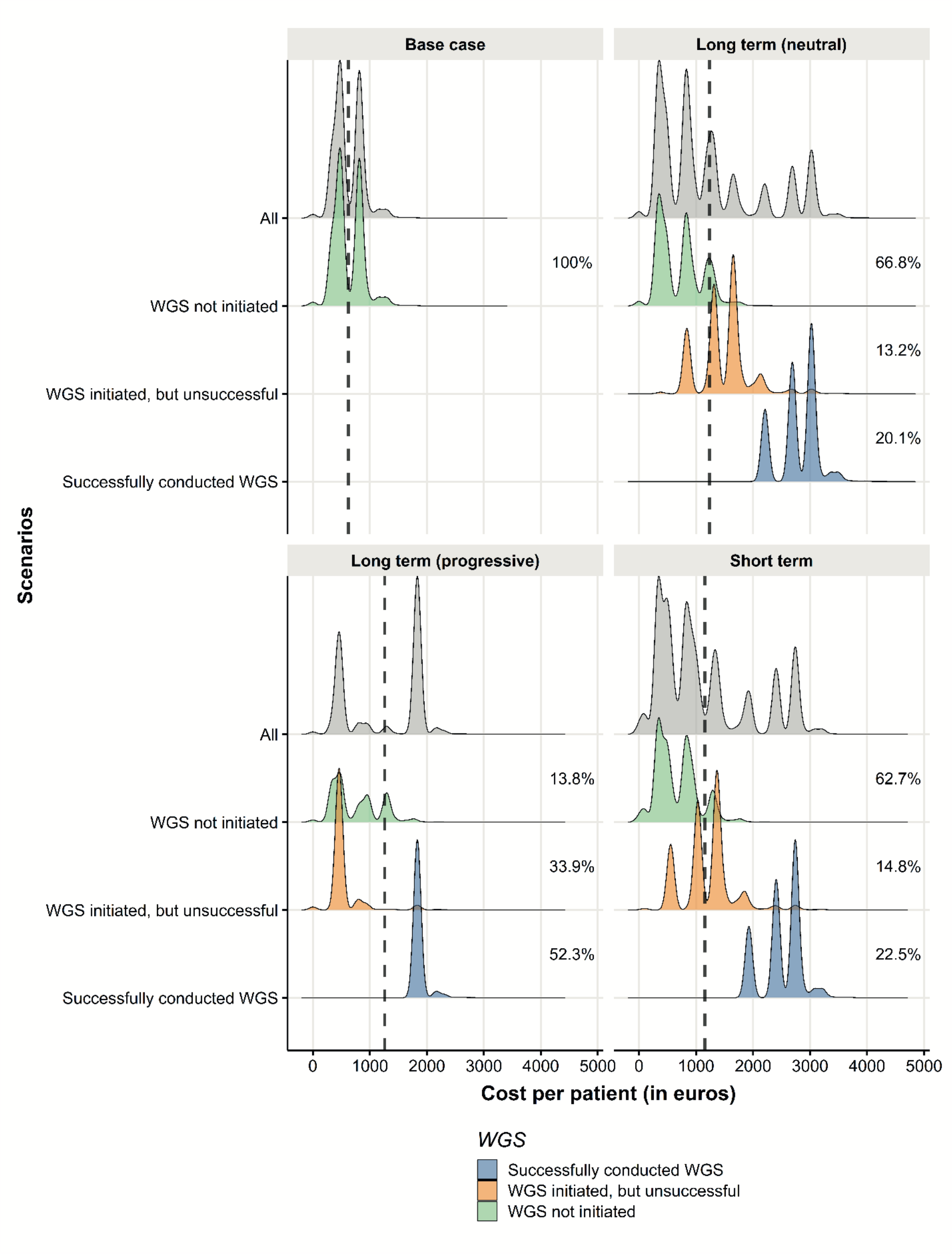
Distribution of the cost per patient for the base case and three scenarios if the cost of consumables were discounted with 50%.

